# Anti-microbial effect of filtered 222nm excimer lamps in a hospital waiting area

**DOI:** 10.1101/2021.09.03.21263096

**Authors:** Jacob Thyrsted, Søren Helbo Skaarup, Andreas Fløe Hvass, Sara Moeslund Joensen, Stine Y. Nielsen, Elisabeth Bendstrup, Pernille Hauschildt, Christian K. Holm

**Affiliations:** Department of Biomedicine, Aarhus University, Aarhus, Denmark; Deparment of Respiratory Diseases and Allergy, Aarhus University Hospital, Skejby, Denmark; Department of Clinical Microbiology, Lillebaelt Hospital, Vejle, Denmark

**Author notes:** Correspondence to: Christian K. Holm, Pernille Hauschildt. These authors contributed equally (Søren Helbo Skaarup, Andreas Fløe Hvass).

**Keywords:** Hospital acquired infections, 222nm excimer, Krypton Chloride (KrCl), Ultraviolet light, Respiratory infections, Immunocompromised state

## Abstract

**Background:** Hospital aquired infections is a considerable challenge for vulnerable patients. Ultraviolet light based on the excitation of mercury emit light at 254nm and has well established antimicrobial effects but the use hereof in populated areas is hindered by the carcinogenic properties of 254nm. This is in contrast to the recently developed excimer lamps based on krypton chloride (KrCl). These lamps emit light with a peak intensity at a wavelength of 222nm and have recently been demonstrated to have broad bactericidal and viricidal effects including efficient inactivation of SARS-CoV2. It is, however, unclear how efficiently 222nm lamps perform in a real-life setting such as a hospital waiting area. In this study we aimed to assess the antimicrobial efficacy of filtered 222nm excimer lamps in a real-world setting at an out-patient pulmonology clinic.

**Methods:** Filtered KrCl 222nm excimer lamps (UV222 lamps) were installed in a densely populated waiting room at the out-patient waiting area at Department of Respiratory Diseases and Allergy at Aarhus University Hospital, Aarhus, Denmark. Furniture sufaces were sampled and analyzed for bacterial load in a single arm interventional longitudinal study with and without exposure to filtered 222nm UVC-light. Furthermore, bacterial species were identified using MALDI-ToF mass-spectrometry.

**Findings:** The exposure to filtered 222nm UVC-light significantly reduced the number of colony-forming-units, and patches with high desity of bacteria. Pathogenic bacteria such as *Staphylococcus Aureus* and *Staphylococcus Epidermidis* were detected only in the non-exposed areas suggesting that these species are highly sensitive to inactivation by 222nm UVC-light.

**Conclusion:** Filtered 222nm UVC-light is highly anti-microbial in a real-world clinical setting reducing bacterial load and eradicating clinically relevant bacteria species. Filtered 222nm UVC-light has the potential to become an important part of current and future anti-microbial prevention in the clinic.

## INTRODUCTION

Hospital-acquired infections (HAI) is a major health problem on a global scale; it is estimated that 23.6% of hospital-treated sepsis cases are hospital acquired (Markwart, Saito et al, Intensive Care Med 2020). In the USA alone, the applied cost for health care providers from HAI has been estimated to consist up to 14.9 billion USD in 2016 (forrester, Maggio et al, j patient saf 2021), and furthermore, HAI account for up to 37,000 annual deaths in Europe and 99,000 annual deaths in the USA (WHO). The recent Sars-CoV-2 pandemic has highlighted our vulnerability toward spread of highly contagious infectious agents, and the importance of implementing and further developing techniques to prevent spread of contagions, especially in places where large numbers of persons pass by, or where particularly vulnerable persons gather. The clinic of pulmonology is an example of such a high-risk area. Here, numerous patients with respiratory diseases are crowed in a small area. Some patients visit the clinic to commence or follow-up treatment for infectious respiratory diseases such as complicated pneumonia, lung absces, fungal pneumonia, tuberculosis or bronchiectasis and wait near others patients who are on immunosuppressive therapy, have had lung transplantation or have severely reduced lung function. Thus, spreading infection from one patient to another may have dramatic consequences in this population. Use of anti- microbial measures such as frequent cleaning of rooms and furniture along with separation of patients and focus to reduce waiting time may lower risk for infections, but more efforts to modify the risk are needed and advanced techniques have evolved in recent years to supplement the effect of such measures.

Ultraviolet (UV) light is well-known to possess excellent disinfecting properties, by inducing the formation of pyrimidine dimers in RNA and DNA, thereby interfering with transcription and replication(1, 2). “Classical” 254 nm UV light has long been used in biological safety cabinets in laboratories. However, the fact that conventional 254 nm UV light is highly carcinogenic in humans(3) limits its potential for anti-septic utilization in locations with high risk of person-to-person transmission of contagions. Recently, a new generation of filtered excimer lamps based on excitation of krypton chloride (KrCl), generating 222 nm UVC light, has been introduced. In contrast to UVC light at 254nm, filtered 222nm excimer lamps (UV222 lamps) can be safely installed in populated areas (4-11).

In order to test the anti- microbial potential of UV222 lamps to reduce the risk of nosocomial spread of infections, we tested the effect of installing filtered UV222 lamps in a out-patient clinic waiting room on bacterial load on exposed surfaces.

## METHODS

### Aims, setting and study design

The primary aim of this study was to investigate the anti-microbial potential of UV222 lamps in an out-patient hospital setting. A prospective longitudinal single arm interventional study with serial sampling was designed and set up in the waiting area at the out-patient clinic at the Department of Respiratory Diseases and Allergy, Aarhus University Hospital, Aarhus, Denmark.

Secondary aims were to evaluate how the UV222 lamps affected bacterial load in the UV222 exposed area compared to chairs placed in the more distant area of the waiting area and thus outside the range of the UV222 lamps. Furthermore, we aimed to identify the bacteria load and determine which bacterial species were present on non-UV-exposed chairs compared to UV222-exposed chairs.

### The waiting area

In the clinics waiting area patients awaits consultation or respiratory examinations. The clinic has services for patients with complicated infectious respiratory diseases, follow up after lung transplantation, severe asthma, interstitial pneumonias, and suspected lung cancer and is equipped with 10 chairs.

### UV222 lamp

The far-UVC source used in this study was a germicidal lamp (UV222, UVmedico, Denmark) based on a filtered KrCl* excimer light source emitting at 222 nm (Care222, Ushio, Japan). The optical filter blocked the remnants in the 230-350 nm emission range which are naturally present in KrCl* excimer lamps. At 222 nm the lamp had a total output of 120 mW, with a full-width half-max emission angle of 60 degrees, resulting in an irradiance of 13.7 μW/cm^2^ at 1 m distance. The output power, optical spectrum and spatial distribution of the lamp has been characterized using a UV calibrated goniometer (“LabSpion” from “Viso Systems”). Input information into the UV222 software contained data on distance to nearest unprotected eye, maximum occupancy time per patient or staff, and dose delivered (dd) per on-off cycle in the farthest distance from the lamp. In this instance, dd for chair seats were 400μJ, which is the approximate equivalence of the dose needed to reduce infectivity of SARS-CoV2 with approximately 90%(4, 12).

### Simulation of waiting room

For simulation and visualisation of the light distribution and energy levels on different surfaces in a room, the DiaLux EVO version 9.2 Light simulation software was used. The fixture files used in the program were verified and measured using the reference LabSpion system and goniometer from VisoSystems. To illustrate the setup and to calculate UV222 doses, a 3D-model of the waiting area and the placement of the lamps was generated (fig. 1a). Evaluation of the overall exposure intensity (fig. 1b) and the exact delivered energy in μW/cm^2^ on each section of the chairs (fig. 1c) was done using this model. Based on the energy values on the outer part of the two chairs positioned under one of the lamps (1,3-2,0 μW/cm^2^), we adjusted the UV222 devices to deliver a total of 400 μJ for each on-cycle. The rationale for this dosage is that many viruses including SARS-CoV2 are significantly inactivated at this dosage(12). Infectious viruses are present mainly in aerosols in the air and thus closer to the lamp. As the intensity of the emitted light increases with increased proximity to the UV222 lamp, viruses suspended in the air would then receive a sufficient 222nm dose to significantly reduce the risk of transmission in one on-cycle(4, 12). The length of the off-cycle was adjusted so that patients were never exposed to more than 22.3mJ in one visit, which is maximum daily exposure limit set by the Danish government and the European Union.

**Figure 1:**
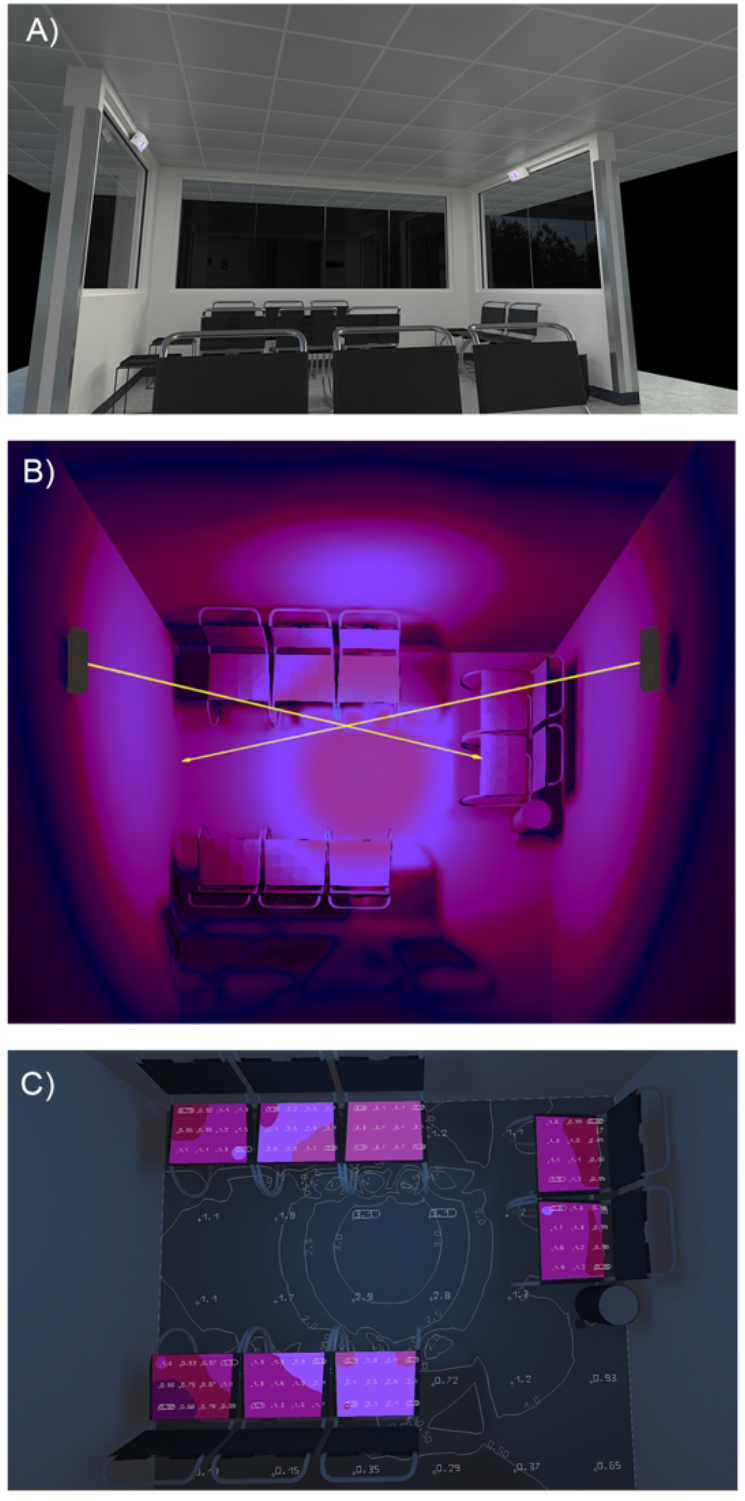
Depiction of how UV222 lamps were installed in a section of the waiting areas at Aarhus University Hospital, Department of Respiratory Diseases and Allergy. A) 3D simulation of the waiting area covered by the UV222 lamps. B) Depiction of 222nm emission from the UV222 lamps. C) Simulation of surface dosage of 222nm in μW.

### Bacterial load sampling

Bacterial loads were determined using Hygicult® TPC sampling kit (Adian) in accordance with the protocol supplied by the manufacturer. The agar-covered sampling sticks were used to collect samples from chair seats and backrests. The sampling sticks were then incubated overnight at 38 deg C before counting bacterial colonies.

Samples were collected at the same time in the afternoon on three consecutive days with the UV222 lamps turned off and repeated for three consecutive days with the UV222 lamps turned on. As the same spots were sampled on each chair on three consecutive days, CFU counts of each day were assessed to see if variance in bacterial load was introduced.

### Maldi-Tof MS identification of bacteria

Matrix-assisted laser desorption ionization time-of-flight mass spectrometry (MALDI-TOF MS), was used to determine which bacterial species were present on non-UV-exposed chairs compared to UV222-exposed chairs. Hygicult® TPC sampling sticks were randomly selected and analyzed.

### Ethical concerns

Following local regulative informed patient consent was not required. No sensitive data was recorded. The UV222 lamp is commercially available and approved by Conformité Euroéenne, the CE-mark, meeting European Union standards for health and safety. Conduction of the study was approved by the chief of department.

### Statistical analysis

Paired students t-test were performed according to analysis of parametric data distribution. A students t-test test was done to evaluate change in bacterial load over the days of sampling. Statistical level of significance was set to 5%. Data were analysed in graphPad Prism.

## RESULTS

### Simulation of 222nm exposure from UV222 lamps installed in the waiting area at Aarhus University Hospital

Using the simulation 3D-model of the waiting room (fig. 1a) the overall exposure intensity (fig. 1b) and the exact exposure on each chair (fig. 1c) was calculated and the UV222 devices were adjusted to deliver a total of 400 μJ for each on-cycle.

### Effect of UV222

Regarding aim 1, we found that UV222 lamps reduced the bacterial load of all chairs as the CFU count was significantly lowered from a mean of 26 CFU pr sample without UV222 exposure to 8 CFU pr sample with UV222 exposure (p<0.0001 by paired t-test) (Fig. 2A). Interestingly, we found the UV222 lamp not only decreased the overall amount of CFU but also efficiently removed all high CFU counts (above 20 CFU per sample). The decrease in CFU was consistent when comparing the sampling days (Fig. 2B-D). Additionally, UV222 exposure was sufficient to decrease the CFU of both seat and backrest of all chairs indicating good coverage of the chairs with 222nm exposure (Fig. 2E-F). Furthermore, we found no pattern of variance throughout the sampling period thus suggesting that the sampling itself did not affect CFU numbers (Fig. 2G).

**Figure 2:**
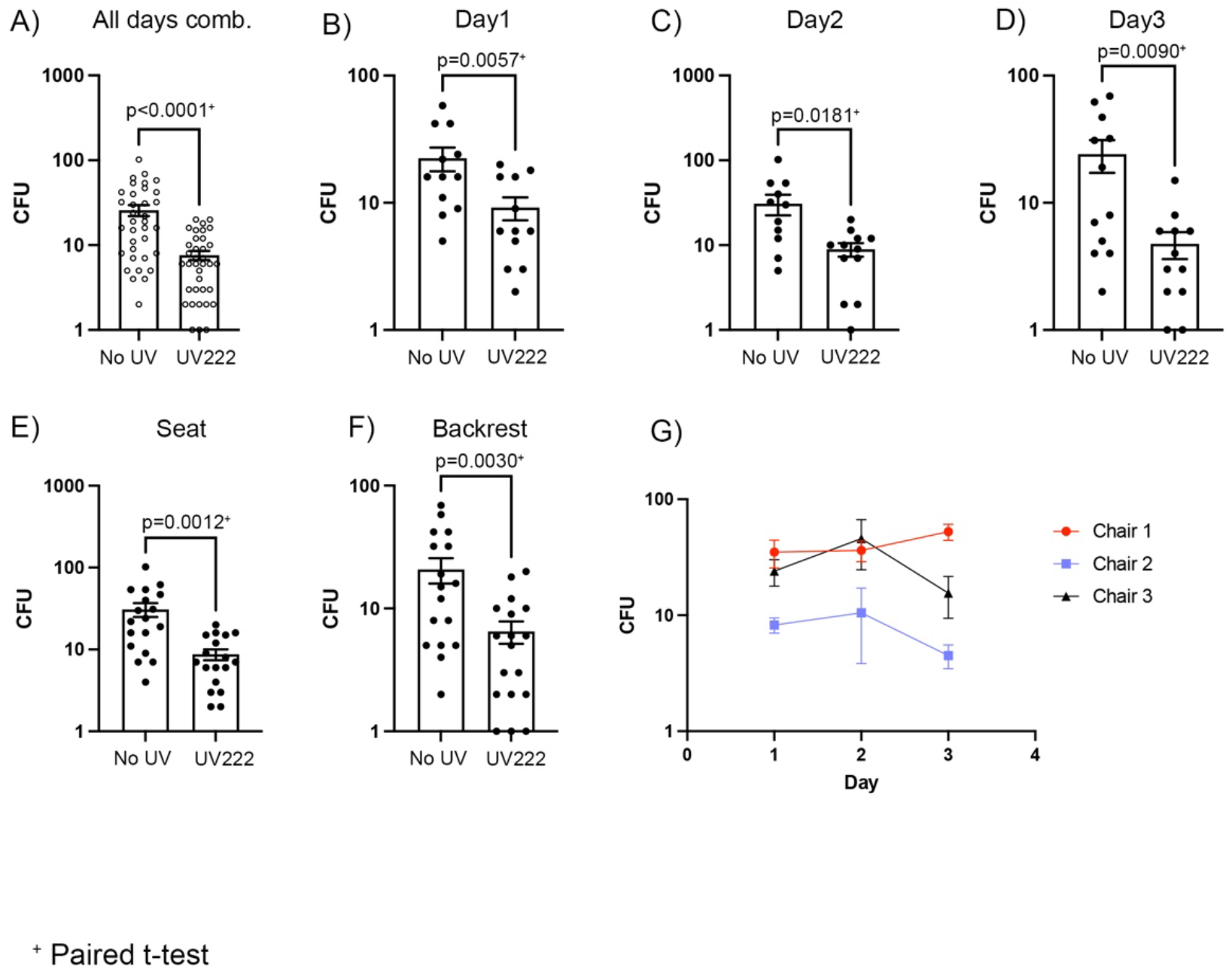
UV222 lamps reduce bacterial load in hospital waiting area. UV222 lamps placed as indicated in figure 1. Bacterial loads were estimated by sampling of seats and backrest of the chair placed in the patient waiting area. Bacterial samples were collected using Hygicult TPC sampling kits. Sampling was performed on three individual days from three individual chairs before installation of the UV222 lamps (No UV) and then again on three individual days and same chairs after UV222 lamp installation. Bacterial colonies were counted after overnight incubation at 38 deg C. UV222 lamps were installed with software to secure that no patients and no staff were exposed to UV222 doses exceeding 23mJ per working day. Bacterial colonies are depicted as total number of colonies A), each individual day B-D), colonies on seats E) and on backrest F). To determine if sampling in itself affected bacterial load we compare the counts between days from the No UV chairs G). Statistical analysis was performed using paired students t-test and p values are depicted together with each figure panel. Bars indicate mean +/-s.e.m and each dot represents one biological sample.

### Bacterial load outside the UV222 exposed area

In aim 2, we found a decrease in the bacterial load within the UV222 zone as compared to the chairs that were not exposed to the UV222 lamps (Fig. 3A). The reduction in CFU was consistent during all three sampling days yet with some variation to the degree of bacterial removal (Fig 3B-D). In accordance with results from the chairs that were exposed to the UV222 light, we observed that CFU was reduced on both the seat and the backseat of all chairs during the days with lamps turned on (Fig. 3E-F).

**Figure 3.**
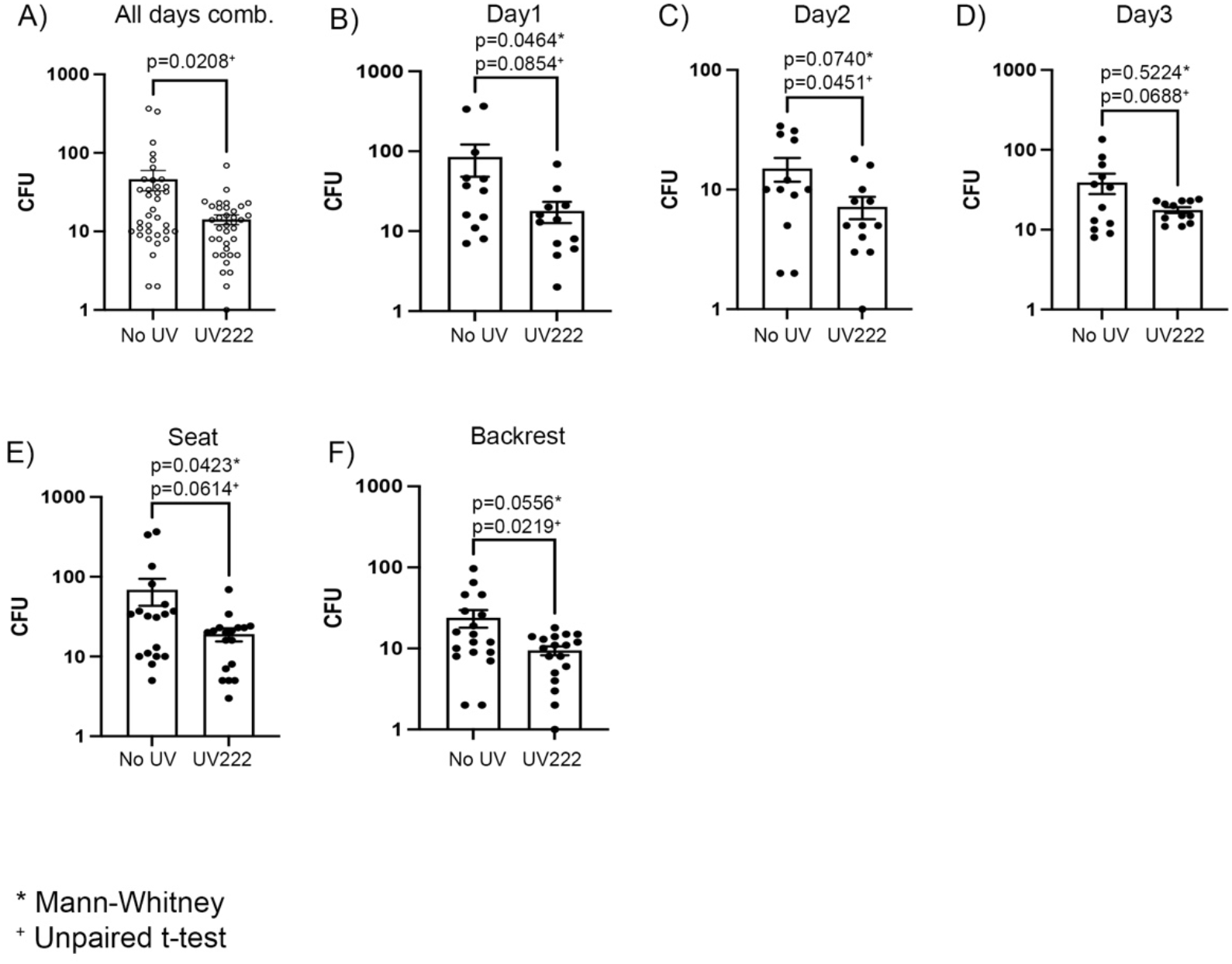
UV222 lamps placed as indicated in figure 1. Bacterial loads were estimated by sampling of seats and backrest of the chairs placed in the patient waiting area. Bacterial samples were collected using Hygicult TPC sampling kits. Sampling was performed on three consecutive days from three individual chairs placed within the UV exposed area (UV222) and from three individual chairs placed outside the UV exposed area (No UV). Bacterial colonies were counted after overnight incubation at 38 deg C. Bacterial colonies are depicted as total number of colonies A), each individual day B-D), colonies on seats E) and on backrest F). Statistical analysis was performed using unpaired students t-test and p values are depicted together with each figure panel. Bars indicate mean +/-s.e.m and each dot represents one biological sample.

### Identification of specific bacterial species using MALDI-TOF

MALDI-TOF MS identified multiple bacterial species. On non-UV chairs 17 different species were identified of which 10 were unique to these chairs as they were not found on UV222 exposed chairs. Interestingly, only 10 different species were found on chairs exposed to UV222, of which seven were seen on both non-UV and UV222 chairs. (Table 1).

**Table 1:** List of identified bacterial species on chairs without UV and with UV. Bacteria names written in **bold** represents species that were only found in one of the conditions.

| Species | No UV | UV222 |
| --- | --- | --- |
| *Acinetobacter Lwoffii* | *Detected* | *Detected* |
| ***Actionocoralia Libanotica*** | ***Not detected*** | ***Detected*** |
| ***Bacillus Cereus*** | ***Detected*** | ***Not Detected*** |
| ***Bacillus Licheniformis*** | ***Detected*** | ***Not detected*** |
| *Bacillus Muralis* | *Detected* | *Detected* |
| *Bacillus Pumilus* | *Detected* | *Detected* |
| ***Bacillus Simplex*** | ***Not detected*** | ***Detected*** |
| *Bacillus Subtilis* | *Detected* | *Detected* |
| ***Brevibacillus Parabrevis*** | ***Detected*** | ***Not detected*** |
| ***Lactobacillus Plantanum*** | ***Detected*** | ***Not detected*** |
| ***Micrococcus Luteus*** | ***Detected*** | ***Not detected*** |
| ***Moraxella Atlantae*** | ***Detected*** | ***Not detected*** |
| *Moraxella Osloensis* | *Detected* | *Detected* |
| ***Staphylococcus Aureus*** | ***Detected*** | ***Not detected*** |
| ***Staphylococcus Capitis*** | ***Detected*** | ***Not detected*** |
| ***Staphylococcus Epidermidis*** | ***Detected*** | ***Not detected*** |
| *Staphylococcus Haemolyticus* | *Detected* | *Detected* |
| *Staphylococcus Hominis* | *Detected* | *Detected* |
| ***Staphylococcus Warneri*** | ***Detected*** | ***Not detected*** |
| ***Staphylococcus Pasteuri*** | ***Not detected*** | ***Detected*** |

## DISCUSSION

This study aimed to evaluate the antimicrobial effect of UV222 light and found that UV222 exposure significantly reduces the overall bacterial load, high CFU counts, and eliminates a number of pathological bacterial species in a waiting area at a pulmonology out-patient clinic.

Conventional mercury UV light has long been known to have disinfecting capability but its carcinogenic potential limits use in areas populated with humans. Searches for a tolerable technique has led to the development of the excimer lamp that generate UV light at 222 nm, a wavelength safe to humans. Most studies published on the antimicrobial efficiency of UV at 222 nm has been performed in laboratory settings as proof of principle(12). While results have been very positive with regards to both human safety and in-vitro antimicrobial effect, this study is the first to show efficacy in a real-life setting and finds two interesting results.

Firstly, the ability of UV222 to significantly decrease the CFU of hospital waiting room chairs highlight the usability of these types of devices in the clinical setting. Of specific interest is the ability of UV222 to eliminate all higher CFU counts possibly stemming from patches of high bacterial density. It is reasonable to assume that these high-density patches potentiate a high-risk bacterial spread. Therefore, removal of all higher bacterial patches could potentially lower spread of bacteria from this surface. This would be of tremendous benefit as a pulmonology out-patient waiting room have a high flow of patients; some patients carry pathogenic and some patients carry antibiotic resistant bacteria and some patients are immunocompromised and may develop fatal illness if exposed to these bacterial patches. Secondly, we show that UV222 remove highly pathogenic bacterial species. Of specific interest is the removal of *Staphylococcus Aureus* as this bacterial species is known for its potential to develop antibiotic resistance. As MRSA is an increasing problem in hospital environments, removal of this species using UV222 could be of high value especially in departments treating patients with compromised immune systems who easily suffer from infections. These patients will also be extra sensitive to *Bacillus Licheniformis, Staphylococcus Epidermidis, Staphylococcus Warneri* and *Micrococcus Luteus* which are all pathogenic in immunosuppressed patients(13). These bacteria were all removed using the UV222 device. The removal of *Bacillus cereus* and *Staphylococcus Capitis* could be of interest to other departments as these bacterial species are known to cause infection following insertion of catheters. Lastly, *Staphylococcus Capitis*, a known producer of biofilm, is also removed using UV222. This could indicate another use of UV222 in removing biofilm producing bacteria. This could be of huge benefit, not only in the clinical world but also in multiple industries suffering from the generation of biofilm.

A limitation of this study is that bacterial samples were only taken from surfaces; no air sampling was done. In patients suffering from respiratory diseases, inhaled bacteria are of special interest, however, it is reasonable to assume that free flowing microbes also are eliminated by UV222 light. Aerosolized particles are exposed from all sides and are more easily eliminated than surface fixed bacteria and are also in closer proximity to the UV222 lamps that receiving a larger dose of 222nm light. A theoretic risk of UV222 could be that elimination of some bacteria allow other species to develop, and if these are pathogenic, the UV222 would potentially create a more dangerous milieu. However, results from the MALDI-ToF rejects this concern. Importantly, the primary study objective was to reduce bacterial load in a clinical setting. A significant reduction was achieved but whether this results in to fewer infections in the patients is yet to be studied.

Finally, following the SARS-CoV-2 pandemic, it has become evident there is a need for better and more efficient disinfection techniques in areas with high density of people. Especially, techniques targeting not only viral pathogens such as SARS-CoV-2 but all disease-causing microorganisms are highly sought after. We did not include SARS-CoV-2 in our study as we expected very low, if any, of this virus in the out-clinic setting as all patients were tested negative before visits. However, vira, including SARS-CoV-2 are more easily eliminated by UV222 than bacteria (Hessling, 2020) and is therefore reasonable to assume that UV222 would inactivae these viruses in this setting as well.

Conclusion

In conclusion, this study shows the usability of UV222 devices in a clinical setting. These devices, being safe for use in areas occupied by humans, has a high potential to become the disinfecting technology of the future as they can generate a continuous anti-microbial environment in areas with high density and high flow of people.

## Data Availability

All data are made available in the manuscript

## Acknowledgements

We would like to acknowledge UVmedico (Aarhus Denmark, UVmedico.com) for supplying filtered 222nm excimer lamps and Christian Byriel for simulation of 222nm exposure in the hospital waiting area.

